# Evaluation of a Large Language Model to Identify Confidential Content in Adolescent Encounter Notes

**DOI:** 10.1101/2023.08.25.23294372

**Authors:** Naveed Rabbani, Conner Brown, Michael Bedgood, Rachel L. Goldstein, Jennifer L. Carlson, Natalie M. Pageler, Keith E. Morse

## Abstract

**Introduction:** In adolescent care, information sharing through patient portals can lead to unintentional disclosures to patients’ guardians around protected health topics such as mental health, sexual health, and substance use. A persistent challenge facing pediatric health systems is configuring systems to withhold confidential information recorded as free text in encounter notes. This study evaluates the accuracy of a proprietary large language model (LLM) in identifying content relating to adolescent confidentiality in such notes.

**Methods:** A random sample of 300 notes were selected from outpatient adolescent encounters performed at an academic pediatric health system. The notes were manually reviewed by a group of pediatricians to identify confidential content. A proprietary LLM, GPT-3.5 (OpenAI, San Francisco, CA), was prompted using a “few-shot learning” method to identify the confidential content within these notes. Two primary outcomes were considered: (1) the ability of the LLM to determine whether a progress note contains confidential content and (2) its ability to identify the specific confidential content within the note.

**Results:** Of the 300 sampled notes, 91 (30%) contained confidential content. The LLM was able to classify whether an adolescent progress note contained confidential content with a sensitivity of 97% (88/91), specificity of 18% (37/209), and positive predictive value of 34% (88/260). Only 40 of the 306 manually reviewed excerpts (13%) were accurately derived from the original note (ie. contained no hallucinations), 22 (7%) of which represented the note’s actual confidential content.

**Discussion:** A proprietary LLM achieved a high sensitivity in classifying whether adolescent encounter notes contain confidential content. However, its low specificity and poor positive predictive value limit its usefulness. Furthermore, an alarmingly high fraction of confidential note excerpts proposed by the model contained hallucinations. In its current form, GPT-3.5 cannot reliably identify confidential content in free-text adolescent progress notes.

## Introduction

In adolescent care, information sharing through patient portals can lead to unintentional disclosures to patients’ guardians around protected health topics like mental health, sexual health, and substance use. Maintaining confidentiality in the wake of the 21st Century Cures Act is crucial to providing equitable care to this population^1^.

Pediatric institutions have employed a variety of methods to prevent such unintentional disclosures, including configuring specific types of medical information from being released to patient portals^2^. A persistent challenge is withholding confidential information recorded as free text in visit notes, up to 21% of which have been shown to contain confidential information^3^. Electronic health records lack functionality to identify and filter such information, and while natural language processing algorithms have shown promise^4^, there is no widely accessible solution.

The emergence of large language models (LLMs) offers a potential solution. LLMs are capable of summarizing large bodies of text and extracting relevant clinical information^5^. The ability of LLMs to accurately identify the presence of confidential content in clinical notes has not been studied. We evaluate the accuracy of a proprietary LLM in identifying content related to mental health, sexual health, and substance use in adolescent progress notes.

## Methods

A random sample of 300 visit notes were selected from outpatient adolescent encounters performed at an academic pediatric health system between January 1, 2016 and December 31, 2019. The notes were manually reviewed by a group of pediatricians specifically trained in adolescent confidentiality laws to identify confidentiality disclosures^3,6^.

A proprietary LLM, GPT-3.5 (OpenAI, San Francisco, CA), was prompted to identify confidential content within these notes. The LLM was accessed via a secure computing platform approved for use with protected health information. Two primary outcomes were considered: (1) the ability of the LLM to determine whether a progress note contains confidential content (note classification); and (2) its ability to identify the specific confidential content within the note (excerpt extraction). For the note classification task, sensitivity, specificity, and positive predictive value were calculated, including 95% confidence intervals assuming a binomial distribution. For excerpt extraction, the model’s proposed excerpts were compared to the original note content to verify accuracy, as LLMs are liable to “hallucinations.” Hallucinations are model outputs that–while plausible–are incorrect or unrelated to the input. Excerpts were also reviewed by a physician to evaluate whether they correctly refer to the actual confidential concepts within the note.

Figure 1 illustrates the structure of the model prompt. A “few-shot learning” approach was used, where the model is given both definitions and examples of confidential content derived from real clinical notes. The prompt is available in the Supplement.

**Figure 1.**
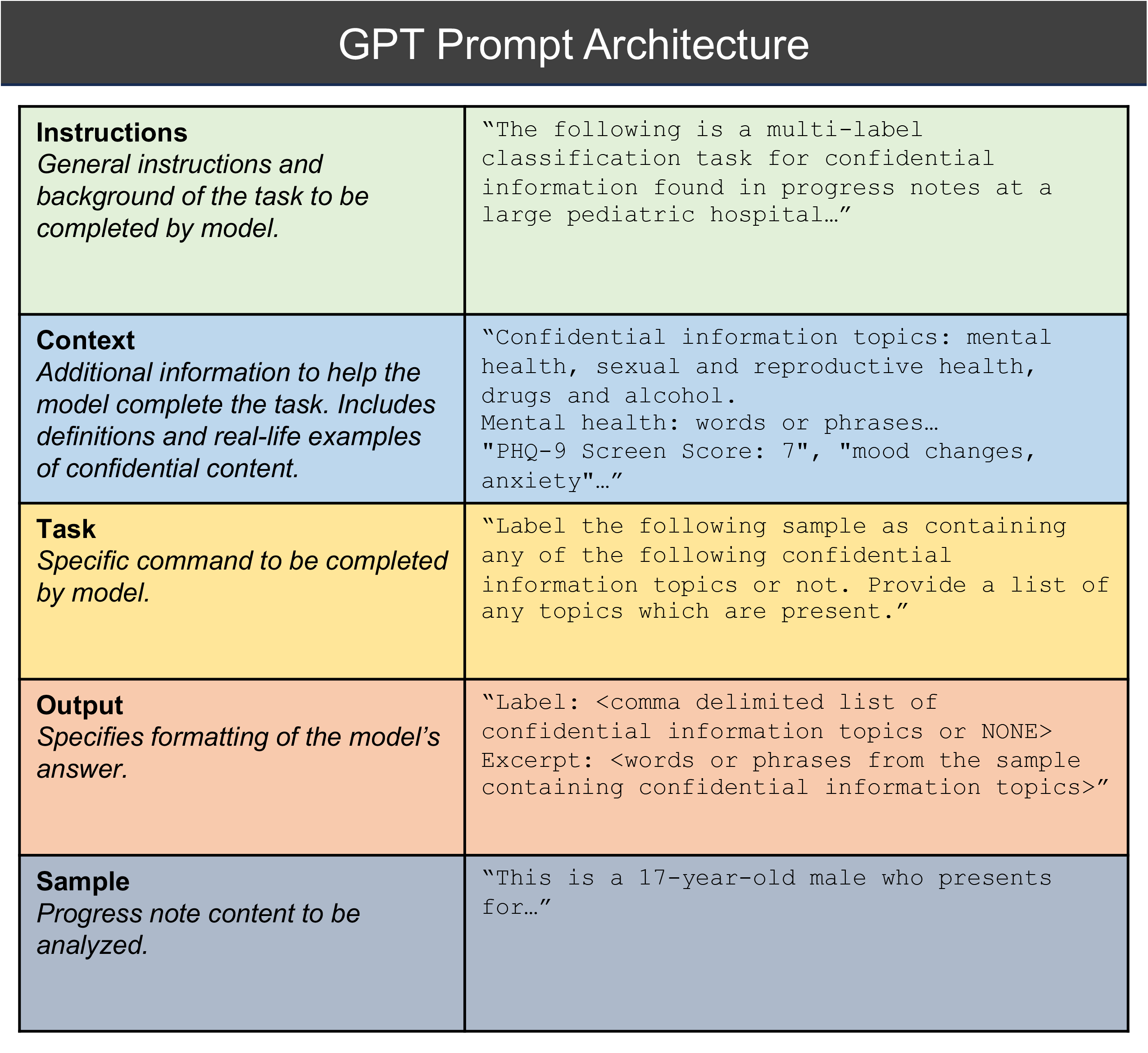
Architecture of the GPT prompt. There are five portions of the prompt provided to the large language model. “Instructions” provide a general description of the problem; “Context” provides the model with further details; “Task” explicitly defines the specific task; “Output” specifies the desired formatting of the model’s response; “Sample” provides the clinical note to be interpreted by the model. The right column shows text from the actual prompt provided to the system.

## Results

Of the 300 sampled notes, 91 (30%) contained confidential content. The LLM was able to classify whether an adolescent progress note contained confidential content with a sensitivity of 97% (88/91), specificity of 18% (37/209), and positive predictive value of 34% (88/260) (Table 1). The LLM proposed a total of 768 confidential excerpts, 306 of which were from a note with confidential content and were reviewed. Only 40 excerpts (13%) were accurately derived from the original note (ie. 87% of excerpts contained a hallucination), and 22 (7%) of excerpts reflected the note’s actual confidential content.

**Table 1.**
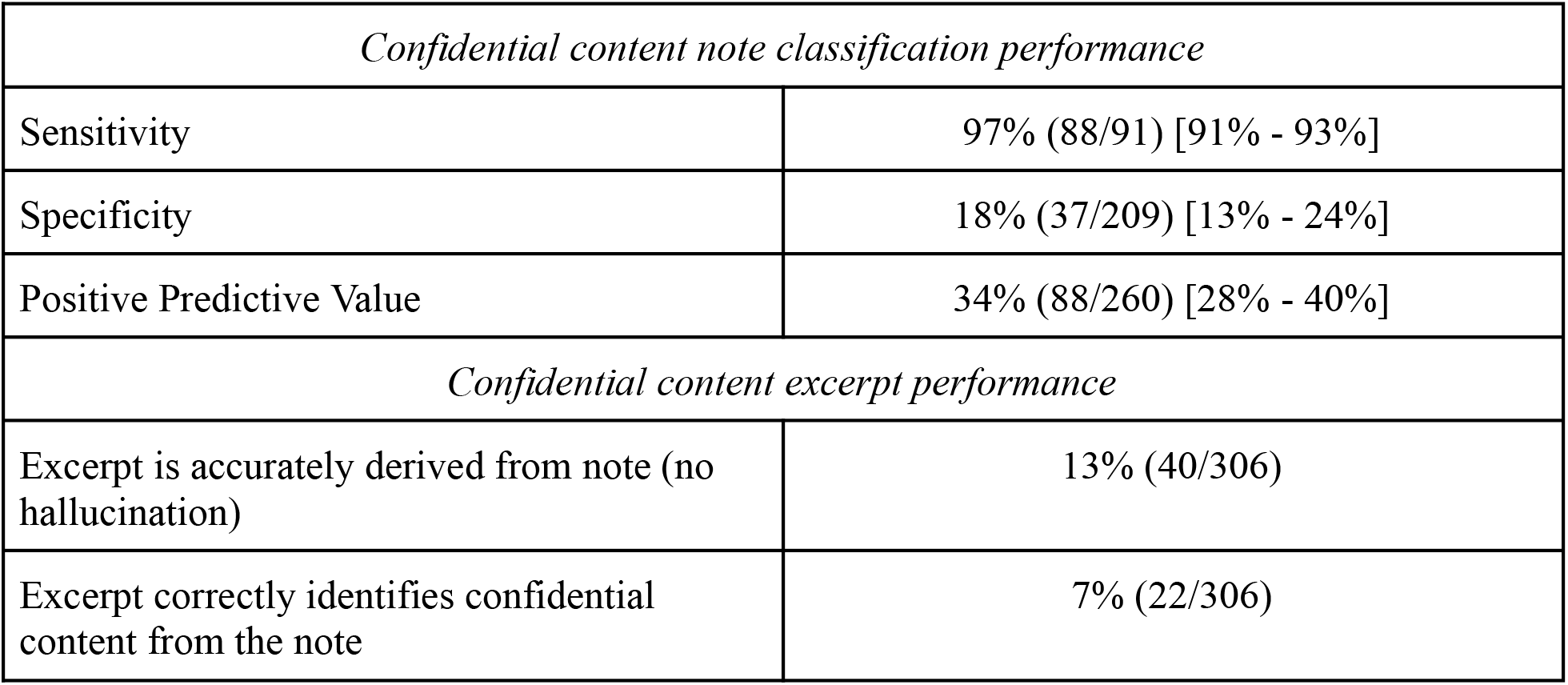
Performance results from GPT-3.5’s ability to identify confidential content in adolescent visit notes. Brackets report 95% confidence intervals.

## Discussion

A proprietary LLM achieved a high sensitivity in classifying whether adolescent notes contain confidential content. However, its low specificity and poor positive predictive value limit its usefulness in hospital operations. Furthermore, the proportion of excerpts containing hallucinations is alarmingly high, such that the output of the model cannot be used for regulatory purposes. With this prompt structure, OpenAI’s GPT-3.5 is not able to reliably identify confidential content in free-text adolescent progress notes.

## Data Availability

The adolescent encounter notes are not publicly accessible as they contain protected health information. However, the model prompt is shared as supplementary material.

## Abbreviations

LLM: Large Language Model

## Contributors Statement Page

Naveed Rabbani: conceptualization, methodology, formal analysis, investigation, data curation, writing - original draft, writing - review & editing, project administration

Conner Brown: methodology, software, data curation, formal analysis, writing - review & editing

Michael Bedgood, Rachel Goldstein, and Jennifer Carlson: methodology, data curation, writing - review & editing

Natalie Pageler: methodology, writing - review & editing

Keith Morse: conceptualization, methodology, investigation, data curation, writing - original draft, writing - review & editing, supervision

All authors approve the final manuscript as submitted and agree to be accountable for all aspects of the work.

