## Supplement for "Evaluation of a Large Language Model to Identify Confidential Content in Adolescent Encounter Notes"

An example of the complete text from the "few-shot learning" prompt used in our GPT-3.5 experiment is provided below. Portions of the prompt that contain identifiable health information, including the encounter note, are redacted in this supplement to maintain patient privacy.

```
### INSTRUCTIONS ###
The following is a multilabel classification task for confidential
information found in progress notes at a large pediatric hospital.
Each sample is a progress note written by the attending provider after a
clinic visit. Some progress notes contain words or phrases which are
considered confidential when the note is shared with a patient's parent
or guardian.
Each note needs to be labeled according to the context provided. It is
important that the task has high precision and excerpts are found in the
sample, so that we can learn more about the words and phrases which are
considered confidential.

### CONTEXT ###
Confidential Information topics: Mental Health, Sexual and Reproductive
Health, Drugs and Alcohol.
Mental Health: words or phrase referring to the mental state of a
patient, the emotional state of a patient or mental health services
provided to a patient. May also refer to child abuse, neglect, emergency
services, mental health services, or shelter services.
Drugs and Alcohol: words or phrase referring to non-prescription drugs,
illegal drugs, alcohol, history of substance use, or services for
substance use. May also refer to drug or alcohol abuse.
Sexual and Reproductive Health: words or phrase referring to the sexual
activity of a patient, services for reproductive health, or medications
for reproductive health. May also refer to pregnancy, contraception,
abortion, rape, sexual assault, intimate partner violence, sexually
transmitted diseases, HIV or AIDS.

"She has a history of anxiety", "PHQ-9 Screen Score: 7", "mood changes,
anxiety, SI/HI","DENIES:depressed mood","no history of suicidal ideation,
depression"
```

"He has no tobacco history on file", "THC, cocaine, Xanax","began after LSD ingestion <redacted>","Patient denies smoking","Alcohol/Drug Use: Yes"
"and she is on OCP", "She continues to use abstinence as her form of contraception","Denies being sexually active","Urine pregnancy test today negative", "Contraception: Abstinence"

### TASK ###
Label the following sample as containing any of the following Confidential Information topics or not. Provide a list of any topics which are present.

### OUTPUT ###
LABEL: <comma delimited list of Confidential Information topics or NONE>
EXCERPT: <words or phrase from sample containing Confidential Information topics>

### SAMPLE ###
<Encounter note text occurs here, redacted due to containing protected health information>
